# Historical Petrol Lead Emissions and Motor Neurone Disease Mortality in Australia

**DOI:** 10.1101/2025.11.06.25339701

**Authors:** Mark A.S. Laidlaw

## Abstract

**Background:** Australian age-standardized Motor Neurone Disease (MND) mortality increased steadily from 1959 and peaked around 2010–2012 and then declined steadily to 2022. The environmental drivers of this trend remain poorly understood. Historical exposure to leaded petrol, reflected in long-term population blood-lead levels, has been proposed as a potential contributor to contemporary MND risk due to the neurotoxicity and long latency associated with lead exposure.

**Methods:** We examined national age-standardised MND mortality in Australia from 1996–2022 in relation to reconstructed cumulative population blood-lead levels derived from digitised Kristensen (2015) data, forward-shifted by 20 years to account for exposure–disease latency. Annual insecticide use per capita was included as a secondary exposure, and calendar year was used to adjust for secular trends. A generalized additive model (GAM) was fitted using a 4-degree-of-freedom spline for cumulative blood-lead levels and linear terms for insecticides and year. Model fit was evaluated using coefficient estimates, joint significance testing for the spline term, and visual inspection of partial-dependence smooths.

**Results:** The GAM R^2^ explained approximately 58.9% (adjusted R^2^ = 49.1%) of year-to-year variation in Australian age-standardised MND mortality rates. The spline for cumulative blood-lead levels was highly significant (p = 0.00024), indicating a strong non-linear association between long-term lead exposure and MND mortality. In contrast, insecticide use showed no statistically meaningful independent effect after adjustment (p = 0.39). Year demonstrated a borderline significant positive association (p = 0.072). Partial-dependence plots revealed substantial curvature in the lead–MND exposure–response relationship, with different historical lead burdens corresponding to distinct changes in predicted MND mortality.

**Conclusions:** These findings support a robust, non-linear association between historical population lead exposure and contemporary MND mortality in Australia, independent of secular trends and insecticide use. The results strengthen the hypothesis that past leaded petrol emissions may contribute to current MND risk, consistent with the long biological persistence and delayed neurotoxic effects of lead. Further work integrating individual-level data, biomarker validation, and mechanistic studies is warranted to clarify causality and quantify population-attributable risk.

**SUMMARY BOX:** 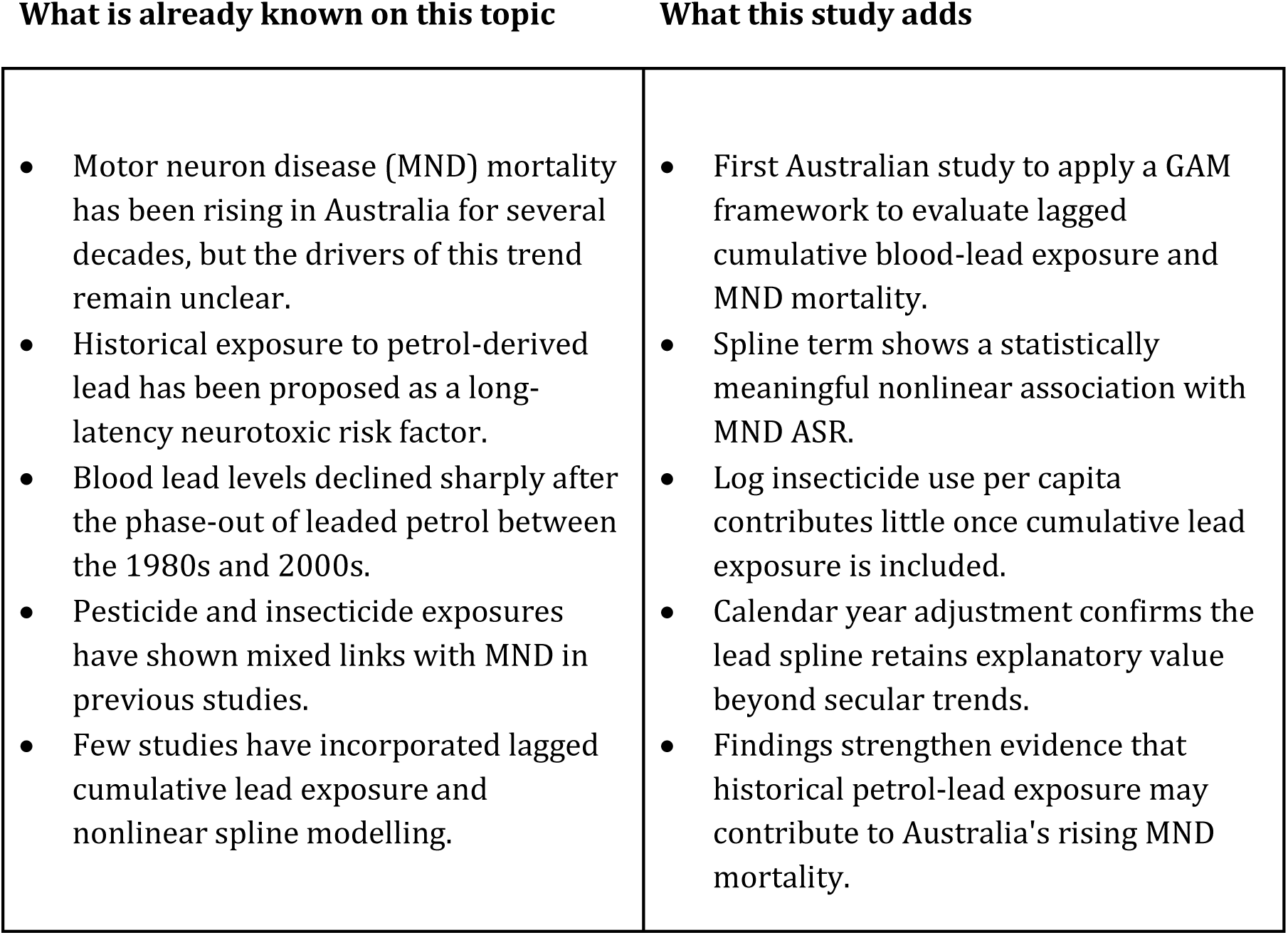

## BACKGROUND

Motor neuron disease is among the most catastrophic neurodegenerative conditions, combining severe healthcare needs with profound personal and family disruption. As the disease progresses, people lose the ability to move, speak, swallow, and breathe, yet their cognitive function generally remains preserved, an experience that can generate considerable psychological strain (Vans et al., 2017). Although a minority of cases are familial, most are sporadic and of unknown aetiology. Increasing evidence implicates environmental toxicants as potential contributors to disease risk, including exposure to heavy metals such as lead (Pb) (Meng et al., 2020; Wang et al., 2014).

Motor neurone disease (also referred to as amyotrophic lateral sclerosis, ALS) carries an unusually high economic and societal cost in Australia. A national cost-of-illness analysis prepared for MND Australia estimated that, in 2015, the total financial impact of the disease was approximately AUD $2.37 billion. This included around $430.9 million in direct expenditure and $1.94 billion attributed to loss of wellbeing and premature mortality, equating to roughly $1.1 million per affected individual (Deloitte Access Economics, 2015). More recent projections commissioned by MND Australia suggest that, without major changes to service delivery, the combined direct and indirect costs of MND will rise to about AUD $5.02 billion by 2025 and could reach AUD $7.51 billion by 2050 (Evohealth, 2025).

Australia provides an advantageous setting to investigate long-term environmental contributions to MND. First, the AIHW maintains a comprehensive mortality dataset with consistent ICD-10 coding, enabling accurate tracking of national trends. Second, detailed historical petrol consumption and lead-content archives allow reconstruction of cumulative petrol-lead emissions and biologically plausible 10–30-year lag intervals (Kristensen, 2015). Third, FAOSTAT (2025) maintains long-term agricultural insecticide usage records, facilitating national exposure reconstruction. Fourth, Australia’s relatively stable population structure and predictable patterns of regulatory change reduce confounding and make secular trends easier to model using temporal smoothers.

## INTRODUCTION

Lee et al., (2026) reported that in Australia, annual deaths attributed to MND rose more than threefold over a 37-year period, increasing from 238 in 1986 to 781 in 2023. Given that only approximately 10 % of MND is understood to be familial, it has been hypothesized that something in the environment must be responsible for the upward trend in MND rates as it is understood that genetic changes could not be responsible for the rise in MND over such a short period of time.

From the early 1930s until its phase-out in the early 2000s, the addition of tetraethyl lead to petrol generated a major anthropogenic source of airborne lead across Australian cities (Kristensen, L.J., 2015) and other industrialized nations such as the United States (Mielke et al., 2011). This contaminated soils in urban centers (Laidlaw and Taylor, 2011; Laidlaw et al., 2014; Taylor et al., 2021; Thorsten et al, 2025). Biomonitoring studies in Australia from the 1970s through to 2002 show that population blood lead levels (BLLs) were substantially higher during the era of widespread leaded petrol use than in the decades following its phase-out (Kristensen, 2015) (Figure 2).

Lead readily crosses the blood–brain barrier, accumulates in bone and neural tissue, and is remobilized with age, providing a biologically plausible mechanism for delayed neurotoxic effects long after exposure has ceased (Hu et al., 1998; Fang et al, 2017; Rueben, 2018). Lead can be measured in bone using portable x-ray fluorescence devices (Specht et al., 2024) or *cumulative* lead exposure can be estimated using blood lead and algorithm-estimated bone lead levels (Wang et al, 2026).

The concept that *cumulative* bone lead levels could be associated with later development of neurodegenerative disorders was assessed by Wang et al. (2026), who found that higher baseline bone lead levels (in the patella and tibia) were significantly associated with an elevated risk of Alzheimer’s disease and all-cause dementia, indicating that long-term cumulative lead exposure is a *key environmental risk factor* for these neurodegenerative disorders.

Lead is now recognized as a potent neurotoxin (Rooney et al, 2012; USEPA, 2024) with the capacity to disrupt mitochondrial function, interfere with neurotransmission, promote oxidative stress, and impair motor neuron integrity in experimental models. Previous ecological research from Australia has reported temporal correlations between historical petrol-lead emissions and contemporary MND mortality rates, suggesting that the disease may reflect long-term consequences of earlier population-level lead exposure (Laidlaw et al., 2015; Zahran et al, 2017).

Despite these observations, several critical limitations remain unaddressed. First, past studies have typically relied on linear regression frameworks, even though the biological effects of cumulative toxicant burden, particularly those with nonlinear dose–response characteristics, are unlikely to be well captured by linear models. Second, few analyses have incorporated lagged cumulative exposure metrics that reflect the protracted latency expected for environmentally mediated neurodegeneration. Third, environmental covariates such as pesticide and insecticide intensity, which have been associated with MND risk in occupational and case–control studies, have not been included in population-level models. Finally, secular changes in disease recognition, diagnostic practices, and population ageing necessitate statistical adjustment for calendar year to avoid attributing long-term mortality trends solely to environmental exposures.

Generalised additive models (GAMs) offer a flexible method for evaluating potential nonlinear relationships between environmental exposures and health outcomes. By incorporating smooth spline functions, GAMs can characterise complex dose–response shapes without imposing restrictive assumptions. In this study, we applied a GAM structure to examine whether a smooth nonlinear function of lagged cumulative blood-lead exposure, reconstructed from historical petrol-lead emissions data, is associated with Australian MND age-standardised mortality rates after adjusting for per-capita insecticide intensity and calendar year.

This approach addresses key methodological gaps in the current literature and provides a more biologically plausible framework for evaluating long-latency neurotoxic exposures. By integrating spline-based exposure reconstruction with temporal covariates, the present study contributes new evidence on the potential environmental determinants of Australia’s rising MND mortality rates.

## METHODS

### Study Design and Outcome Variable

We conducted an ecological time-series analysis using annual Australian national age standardized mortality data for motor neuron disease (MND) from 1986 -2022 (AIHW, 2023). The primary outcome was the age-standardised mortality rate (ASR) for MND (ICD-10 code G12.2). The data for age standardised MND is available from 1959 to 2022 (Laidlaw et al., 2015 and this study), but the age standardised MND data used in the modeling in this study spanned from 1986 to 2022.

### Reconstruction of Historical Lead Exposure

To estimate long-term population exposure to petrol-derived lead, we utilised reconstructed annual BLLs originally digitised from Kristensen (2015) and extended using a shape-preserving monotonic spline (PCHIP). These yearly values were then integrated to generate a cumulative blood lead exposure index, reflecting long-run body burden. A 20-year forward lag was applied to align exposure peaks with the expected latency for environmentally mediated neurodegeneration. The resulting metric was treated as the primary exposure variable.

The optimal lag in MND response was identified by Laidlaw et al., (2015) who previously conducted linear regressions between cumulative petrol-lead emissions (1933–2002) and Australian male and female MND mortality rates, as well as the proportion of MND among all-cause deaths, using forward lags ranging from 10 to 24 years (1962–2013). A 20-year lag provided the best model fit, yielding the highest R² and lowest p-value, and was therefore selected for subsequent analyses of cumulative petrol-lead exposure.

### Insecticide Exposure Variable

Australian yearly national insecticide usage data were obtained from agricultural chemical consumption records (FAOSTAT, 2025). Per-capita usage was calculated by dividing annual national insecticide mass by the corresponding Australian population size (World Bank, 2024). Because raw insecticide values were right-skewed, they were log-transformed and used as a covariate to account for potential confounding by pesticide exposures previously associated with MND risk in occupational studies.

### Temporal Covariate

To adjust for secular changes in diagnosis, reporting practices, population ageing, and unmeasured temporal confounders, the calendar year was included as a linear term (Year) in all models.

### Statistical Modelling: Generalised Additive Model Structure

We modelled the association between MND mortality and cumulative lead exposure using a generalised additive model (GAM)–equivalent spline framework. The model took the form:

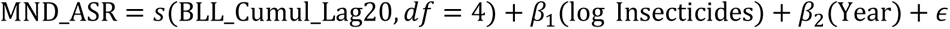

The smooth term s(BLL_Cumul_Lag20) was implemented as a natural cubic regression spline with 4 degrees of freedom, allowing flexible modelling of nonlinear dose–response relationships without overfitting. This approach is statistically equivalent to a GAM smooth term when degrees of freedom (df) is prespecified.

Coefficients for the linear covariates (insecticides and Year) were estimated directly through ordinary least squares (OLS), while the spline basis functions captured nonlinear variation attributable to lagged cumulative lead exposure.

### Model Fitting and Inference

Spline basis functions were generated using the patsy library, and models were estimated using statsmodels OLS.

Spline term significance was evaluated using F-tests derived from the joint contribution of the spline basis coefficients.

### Software

All analyses were conducted in Python (version 3.x) using pandas, statsmodels, patsy, and matplotlib. Spline estimation and plotting were performed using natural cubic splines to replicate mgcv-style GAM behaviour in the presence of SciPy/bspline constraints.

### Ethics and Data Availability

The study used publicly available, aggregate-level environmental and mortality data and therefore did not require human ethics approval. All data used in this analysis are available upon request or can be reproduced directly from the datasets in the supplementary section.

## Results

### Descriptive trends

Australian age-standardized MND mortality increased steadily from 1959 and reached its highest levels around 2010–2012 before showing a modest decline until 2022 (Figure 4). Historical petrol lead exposure in Australia rose sharply after 1958, peaked in the early 1970’s (approximately 8,000 tonnes annually) and declined steadily until it was removed from petrol in 2002 (Figure 1) (Kristensen, 2015). Petrol lead emissions in Australia were approximately 7,000 tonnes per year in 1976 and declined steadily until 2002 when lead in Australian petrol was removed (Figure 2). Petrol lead emissions and the BLL spline were highly correlated during this time-period (r=0.9781, R^2^= 0.9568, p = 1.425e-18) (Kristensen, 2015; Figure 2). This indicates that lead emissions from petrol was the dominant source of lead poisoning in Australia historically. National insecticide use per capita showed a sharp increase from about 1990 to 2000, a moderate undulating increase until about 2017 and declined moderately to 2002 (Figure 4).

**Figure 1.**
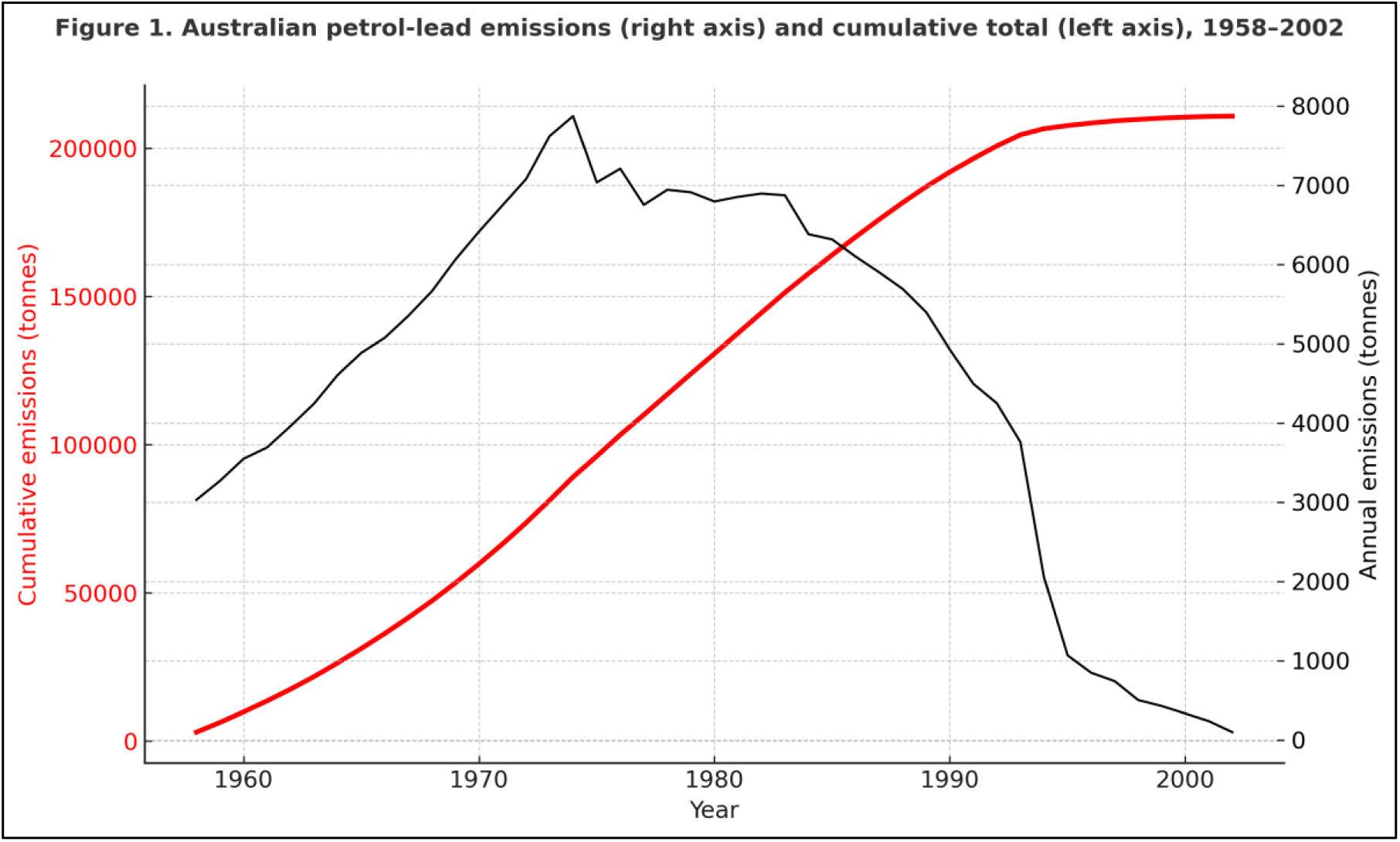
Cumulative Australian petrol lead emissions (tonnes) (1986–2022) and annual petrol-lead emissions (tonnes) (1958–2002). The left y-axis shows cumulative petrol lead emissions (tonnes; thin red line) the right y-axis shows annual petrol-lead emissions (tonnes; thin black line). Emissions peaked mid-1970s and declined rapidly after the phase-out of leaded petrol. The rise in MND mortality follows by about 20 years. *Sources:* AIHW (1986–2022); Kristensen (2015).

**Figure 2.**
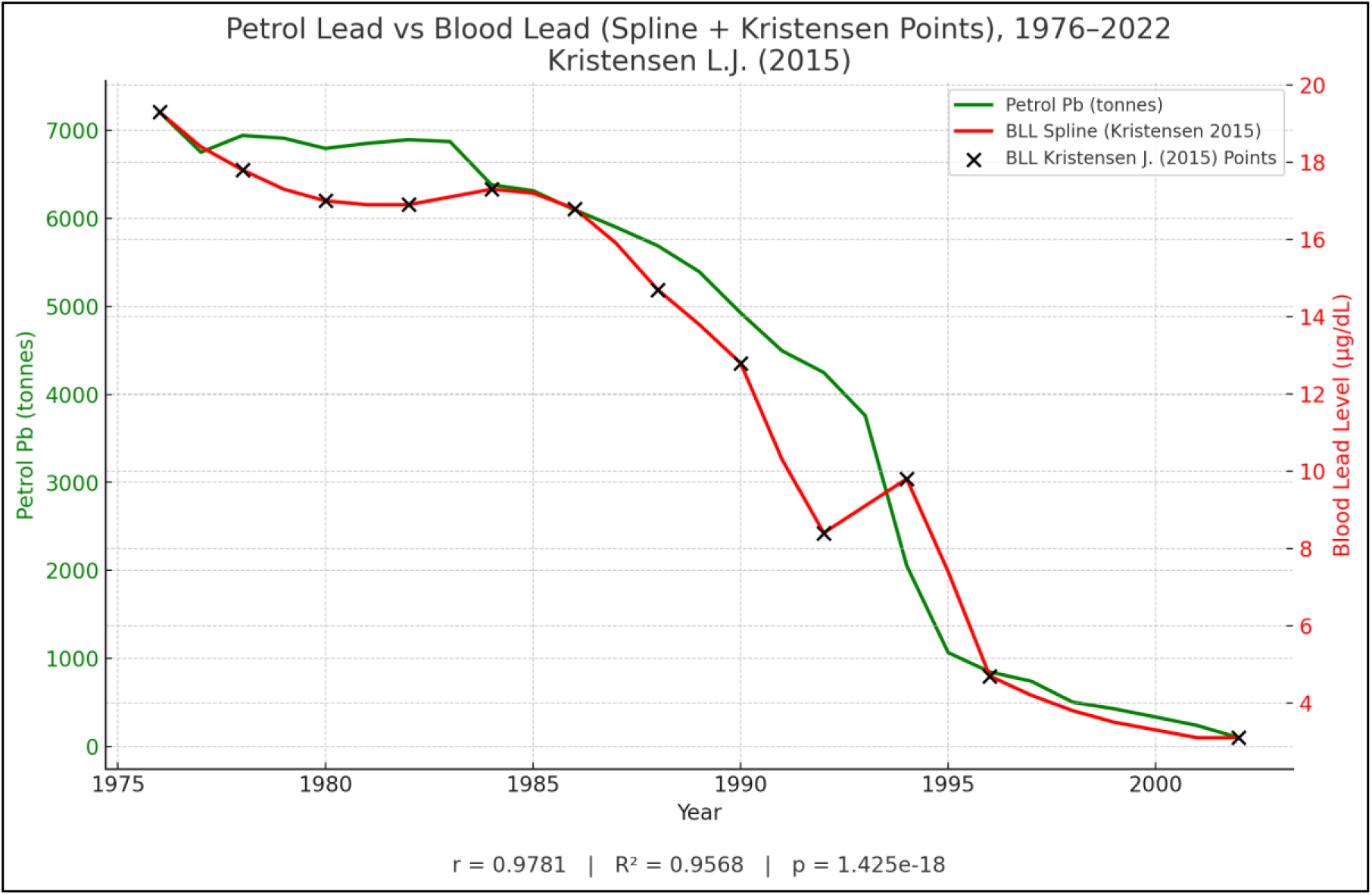
Trends in petrol-lead emissions and reconstructed blood lead levels in Australia, 1976–2022. Annual petrol-derived lead emissions (Au_Petrol_Pb, left Y-axis) are shown alongside reconstructed population BLLs from Kristensen L.J. (2015), including the fitted spline (Au_Bll_Krist_2015_Splin, right Y-axis) and original biomonitoring measurements (Au_Bll_Krist_2015, black points). Petrol-lead emissions declined markedly from the late 1970s through the 1990s, with a parallel reduction in estimated BLLs, reflecting the national phase-out of leaded petrol. The spline-based blood lead estimates were strongly correlated with annual petrol-lead emissions (r = 0.9781; R² = 0.9568; p = 1.425 × 10⁻¹⁸), indicating that reconstructed blood lead trajectories closely tracked historical variation in petrol-lead emissions over this period. (Figure 1).

**Figure 3.**
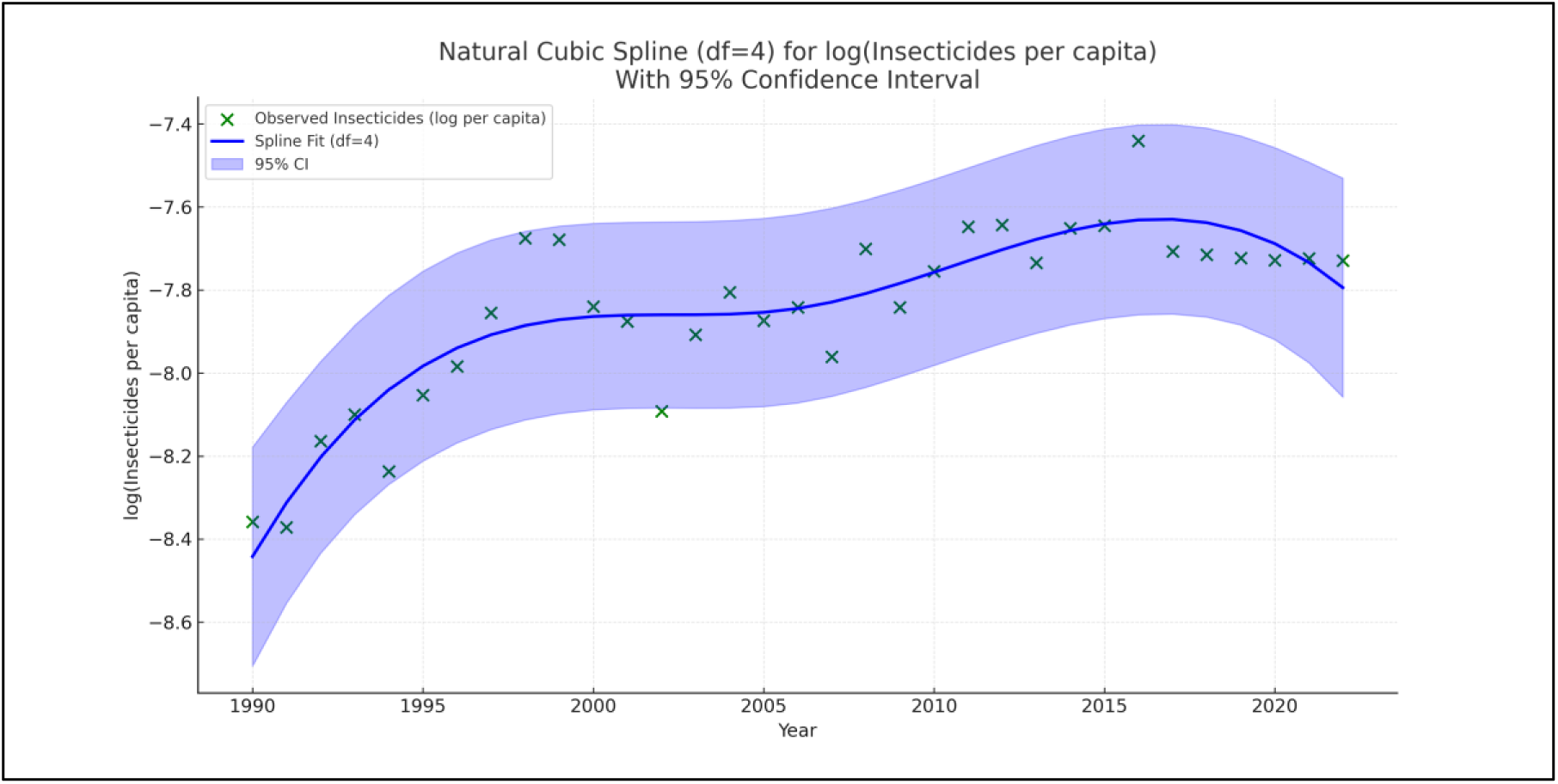
Natural cubic spline (df = 4) fit for log-transformed insecticide use per capita in Australia, with 95% confidence interval (1990–2022). Annual log-transformed insecticide use per capita (Au_log_insecticides_per_capita) is shown with a natural cubic spline fit (df = 4) to characterise long-term nonlinear trends. The shaded region represents the 95% confidence interval of the smoothed estimate. Insecticide intensity increased throughout the 1990s, stabilised during the 2000s, and displayed modest fluctuations in the 2010s, with no sustained upward or downward trend in recent years. This temporal pattern provides context for interpreting the relatively small contribution of insecticide use in adjusted GAM models of national MND mortality.

**Figure 4.**
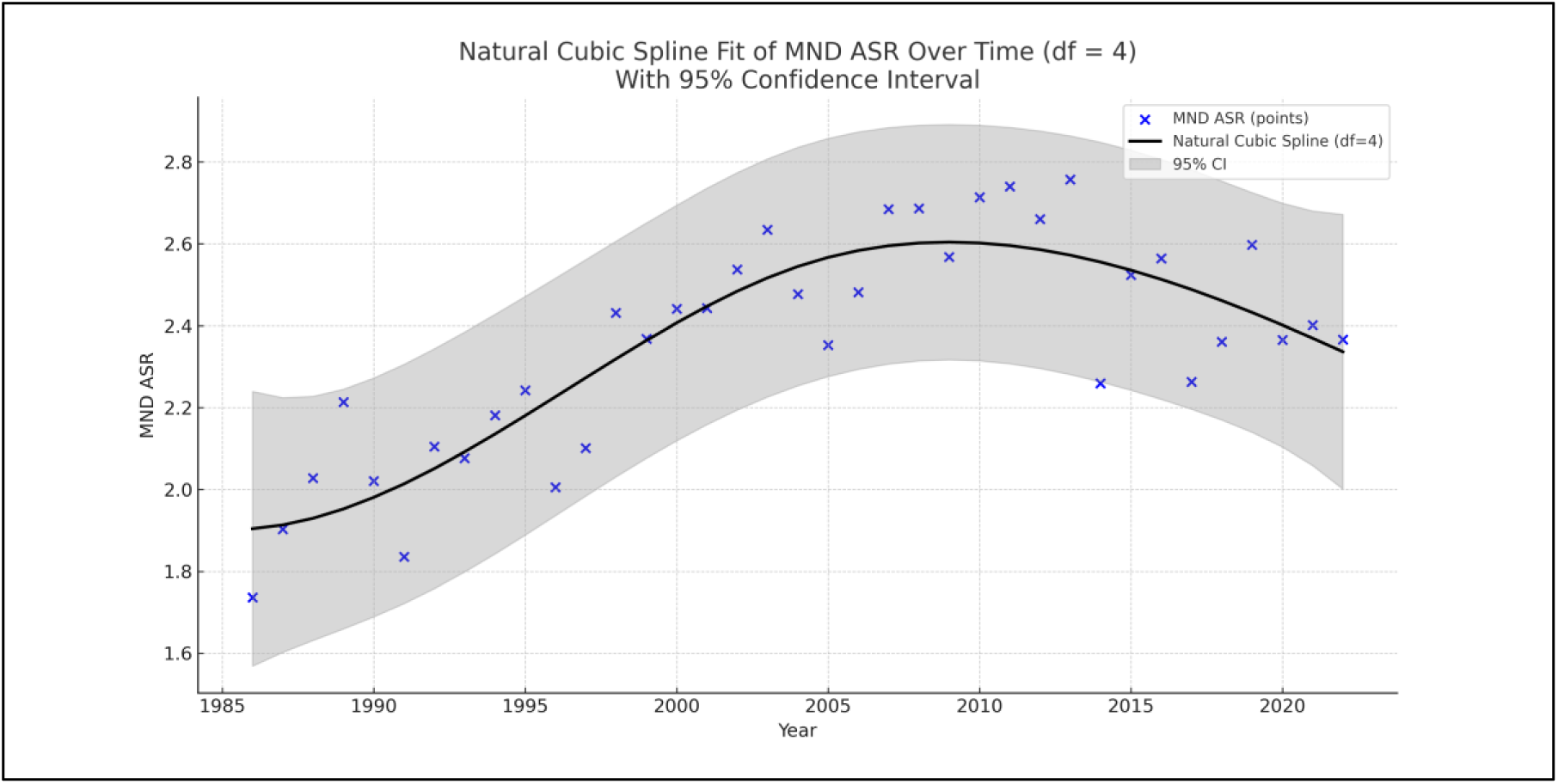
Natural cubic spline fit of national MND age-standardised mortality rates (ASR) with 95% confidence interval, Australia 1986–2022. Annual motor neuron disease (MND) age-standardised mortality rates (*Au_MND_ASR*) are shown with a fitted natural cubic spline (df = 4) to characterise nonlinear temporal trends. The shaded region represents the 95% confidence interval of the spline estimate. MND ASR increased gradually from the late 1980s through the mid-2000s, followed by a modest decline in more recent years. The spline highlights sustained long-term upward drift during the 1990s and early 2000s, consistent with reported increases in national MND mortality over this period.

**Figure 5.**
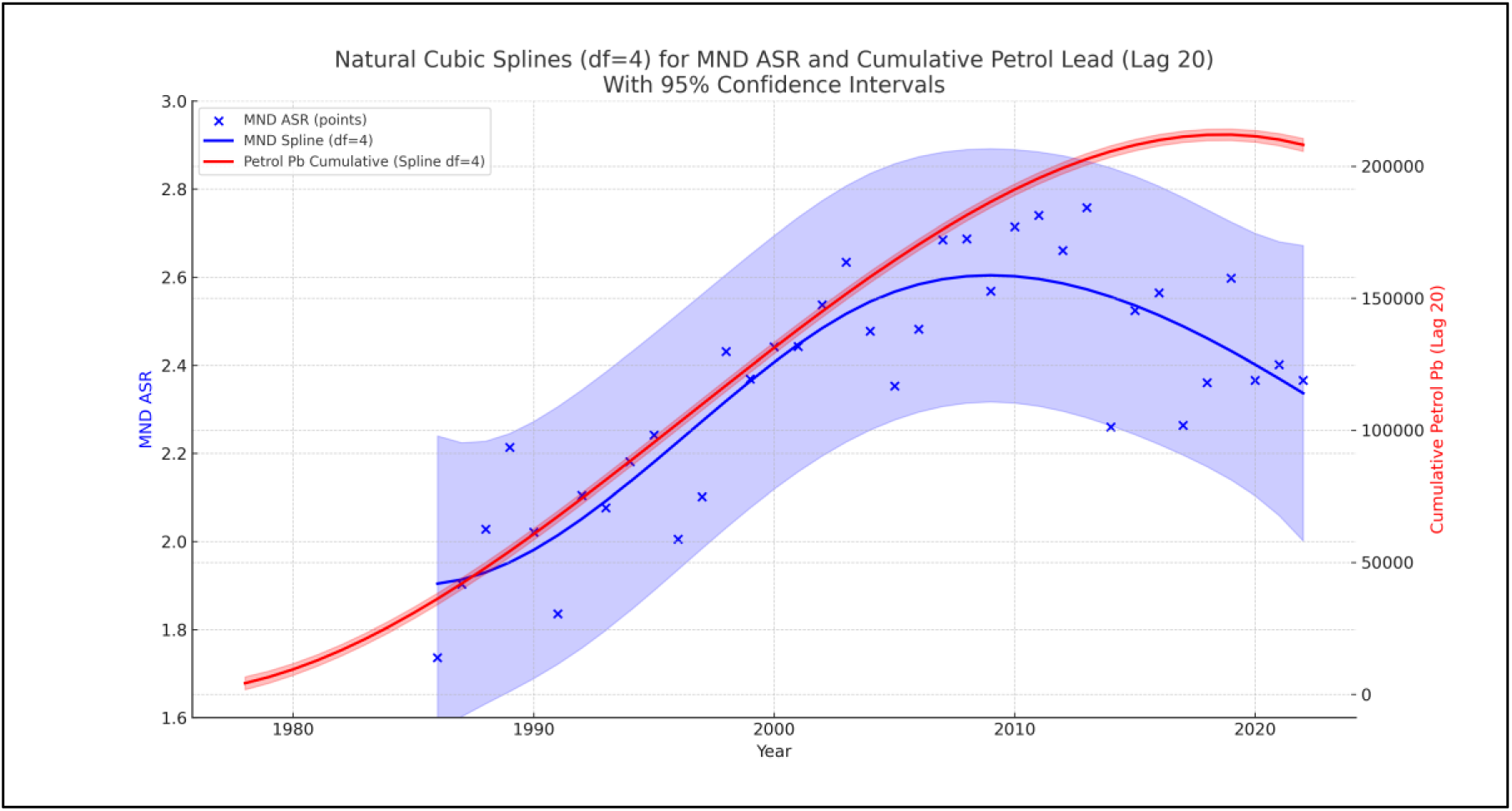
Natural cubic spline fits (df = 4) for national MND age-standardised mortality and cumulative petrol-derived lead exposure (20-year lag), with 95% confidence intervals. (Figure 4) Annual motor neuron disease (MND) age-standardised mortality rates (Au_MND_ASR) are shown alongside the 20-year lagged cumulative petrol-lead exposure (Au_Cumul_Petrol_Pb_Lag_20), with each series fitted using a natural cubic spline (df = 4). Shaded regions represent 95% confidence intervals for each spline. MND mortality rose steadily from the late 1980s through the mid-2000s before stabilising and beginning to decline slightly in more recent years. Lagged cumulative petrol-derived lead exposure follows a similar nonlinear trajectory, increasing sharply throughout the 1980s and 1990s and plateauing after 2010. The overlapping temporal patterns reinforce the plausibility of a long-latency relationship between historical petrol-lead emissions and contemporary MND mortality trends in Australia. (Figure 4)

**Figure 6.**
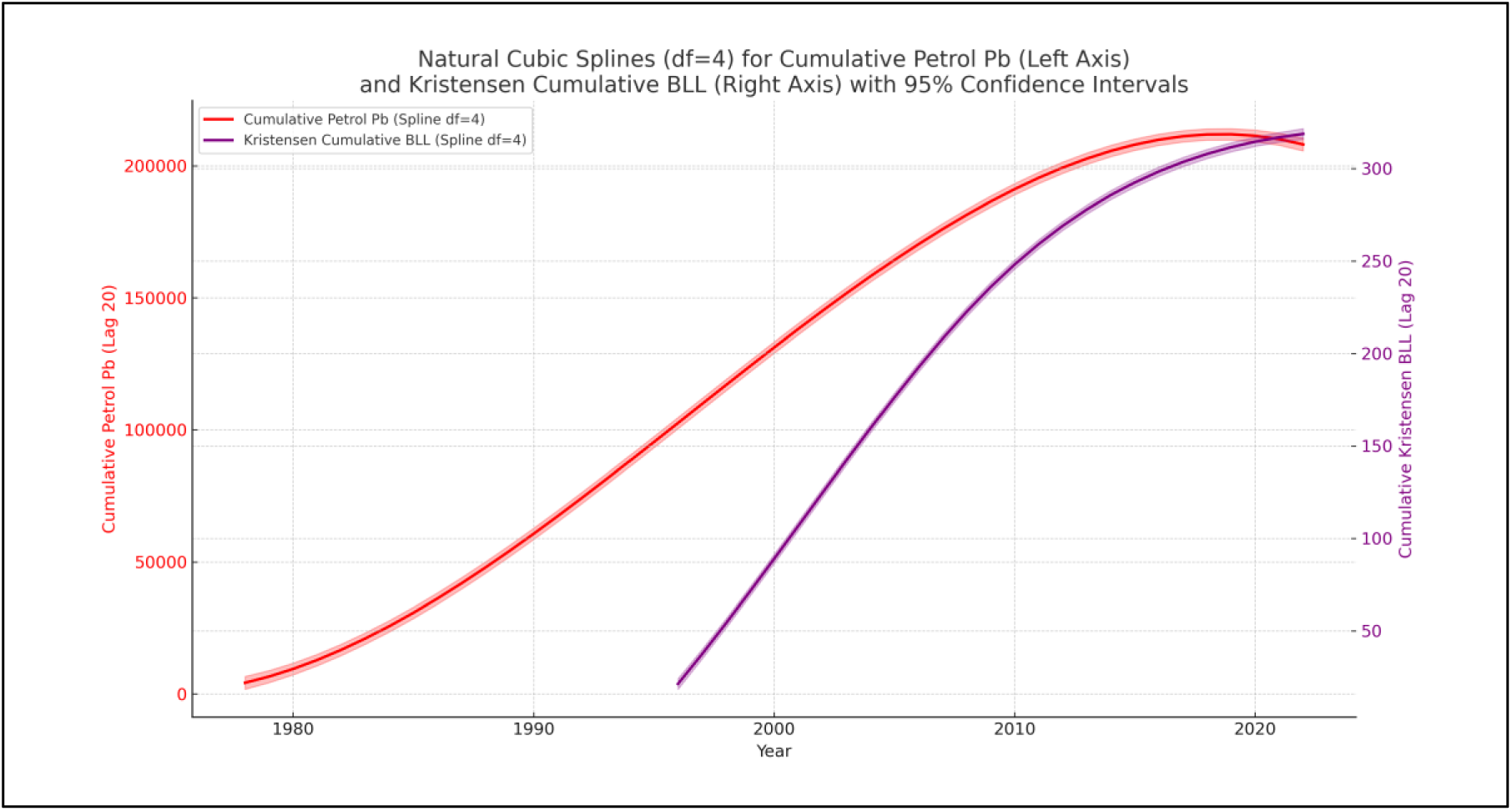
Natural cubic spline trajectories (df = 4) for lagged cumulative petrol-derived lead and cumulative Kristensen blood lead estimates, with 95% confidence intervals. Twenty-year lagged cumulative petrol-derived lead exposure (Au_Cumul_Petrol_Pb_Lag_20, left Y-axis (units = tonnes)) and cumulative reconstructed BLLs from Kristensen L.J. (2015) (Au_Bll_Krist_2015_Cumul_Splin_Lag_20, right Y-axis (units = micrograms per deciliter)) are shown with natural cubic spline fits (df = 4) and associated 95% confidence intervals. Both series demonstrate steep increases during the 1980s and 1990s, reflecting the cumulative legacy of leaded-petrol emissions, followed by a plateauing trend after 2010 as national lead emissions and population BLLs declined. The parallel nonlinear trajectories highlight the close correspondence between cumulative exposure metrics derived from emissions data and reconstructed biomonitoring estimates. (Figure 3) (Figure 4)

To formally evaluate whether long-term historical lead exposure and insecticide use were associated with Australian age standardized MND mortality rates, we fitted generalized additive models (GAMs) to annual age-standardized MND mortality rates over the observation period. Models included a nonlinear spline term for 20-year–lagged cumulative population blood-lead burden, with log insecticide use per capita and calendar year entered as covariates. This approach allowed assessment of both the shape of the exposure–response relationship and the independent contribution of insecticide use after accounting for historical lead exposure and secular trends.

A generalized additive model (GAM) was fitted to Australian annual age-standardized motor neuron disease (MND) mortality rates (1996–2022), including a natural cubic spline (df = 4) for 20-year–lagged cumulative population blood-lead burden, with log-transformed insecticide use per capita and calendar year included as linear covariates. In the full adjusted model, the spline term for lagged cumulative blood lead was statistically significant on joint testing (p = 0.00024), indicating a robust nonlinear association between historical population lead exposure and contemporary MND mortality. The fitted model explained approximately 58.9% of year-to-year variation in national MND age-standardized MND mortality rates (adjusted model fit = 49.1%). In contrast, log insecticide use per capita was not independently associated with MND mortality after adjustment for cumulative blood lead and year (p = 0.39). Calendar year showed a borderline positive association with MND mortality (p = 0.072).

**Table 1.**
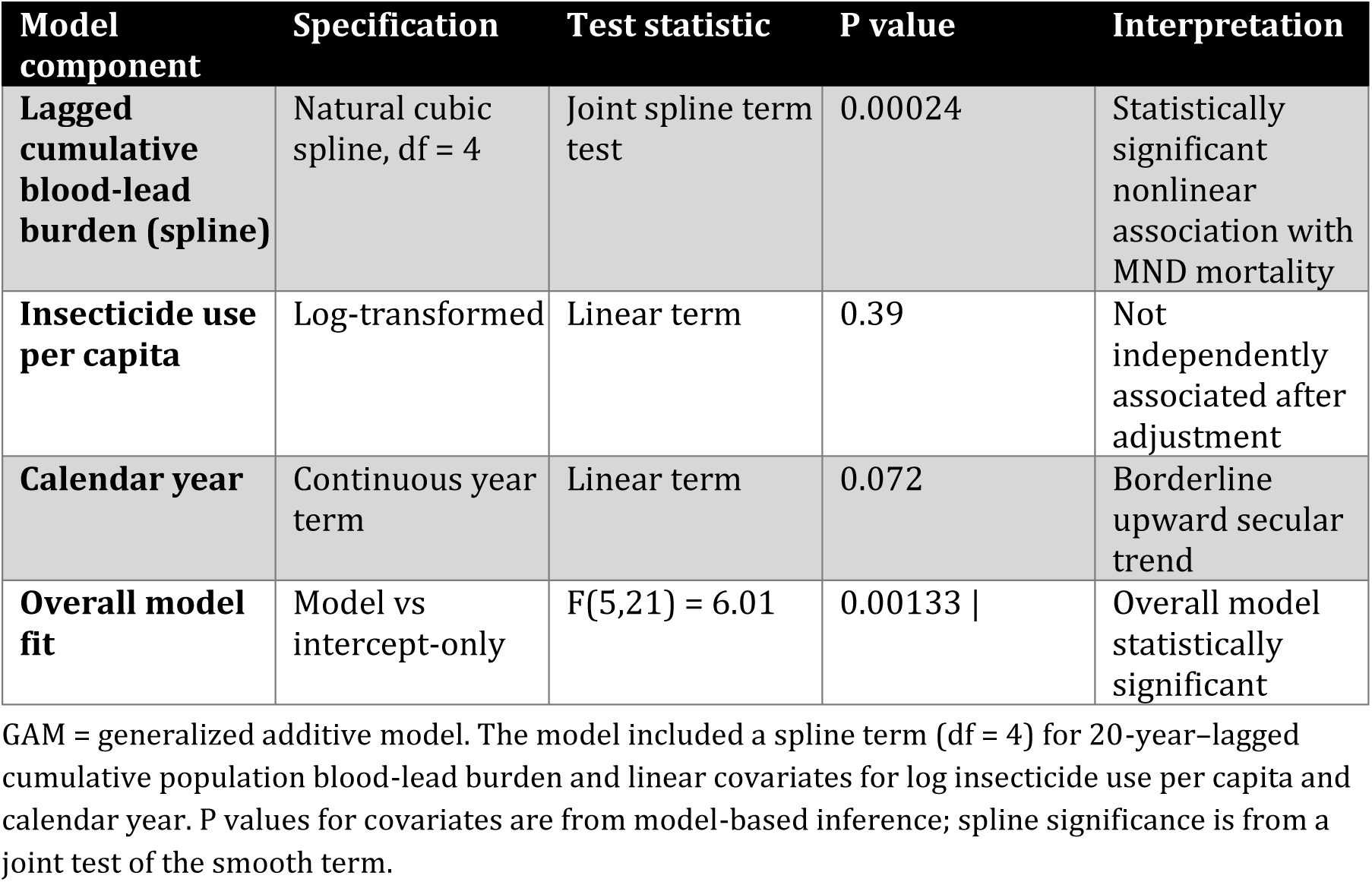
Generalized additive model (GAM) results for Australian age-standardized MND mortality (1996–2022)

| Model component | Specification | Test statistic | P value | Interpretation |
| --- | --- | --- | --- | --- |
| **Lagged cumulative blood-lead burden (spline)** | Natural cubic spline, df = 4 | Joint spline term test | 0.00024 | Statistically significant nonlinear association with MND mortality |
| **Insecticide use per capita** | Log-transformed | Linear term | 0.39 | Not independently associated after adjustment |
| **Calendar year** | Continuous year term | Linear term | 0.072 | Borderline upward secular trend |
| **Overall model fit** | Model vs intercept-only | $F(5,21) = 6.01$ | 0.00133 \| | Overall model statistically significant |

GAM = generalized additive model. The model included a spline term (df = 4) for 20-year-lagged cumulative population blood-lead burden and linear covariates for log insecticide use per capita and calendar year. P values for covariates are from model-based inference; spline significance is from a joint test of the smooth term.

### GAM spline term for lagged cumulative blood lead

The natural cubic spline term (df = 4) for lagged cumulative blood lead demonstrated a statistically meaningful and nonlinear association with age standardized MND mortality. Examination of the spline function revealed an initially shallow slope at lower cumulative exposure levels, followed by a steeper rise across the mid-range of the exposure distribution (Figure 7). This pattern indicates that the association between cumulative lead burden and age-standardized MND mortality is not purely linear, and that increases in cumulative lead exposure correspond to disproportionately higher age-standardized MND ASR values at certain exposure thresholds.

**Figure 7.**
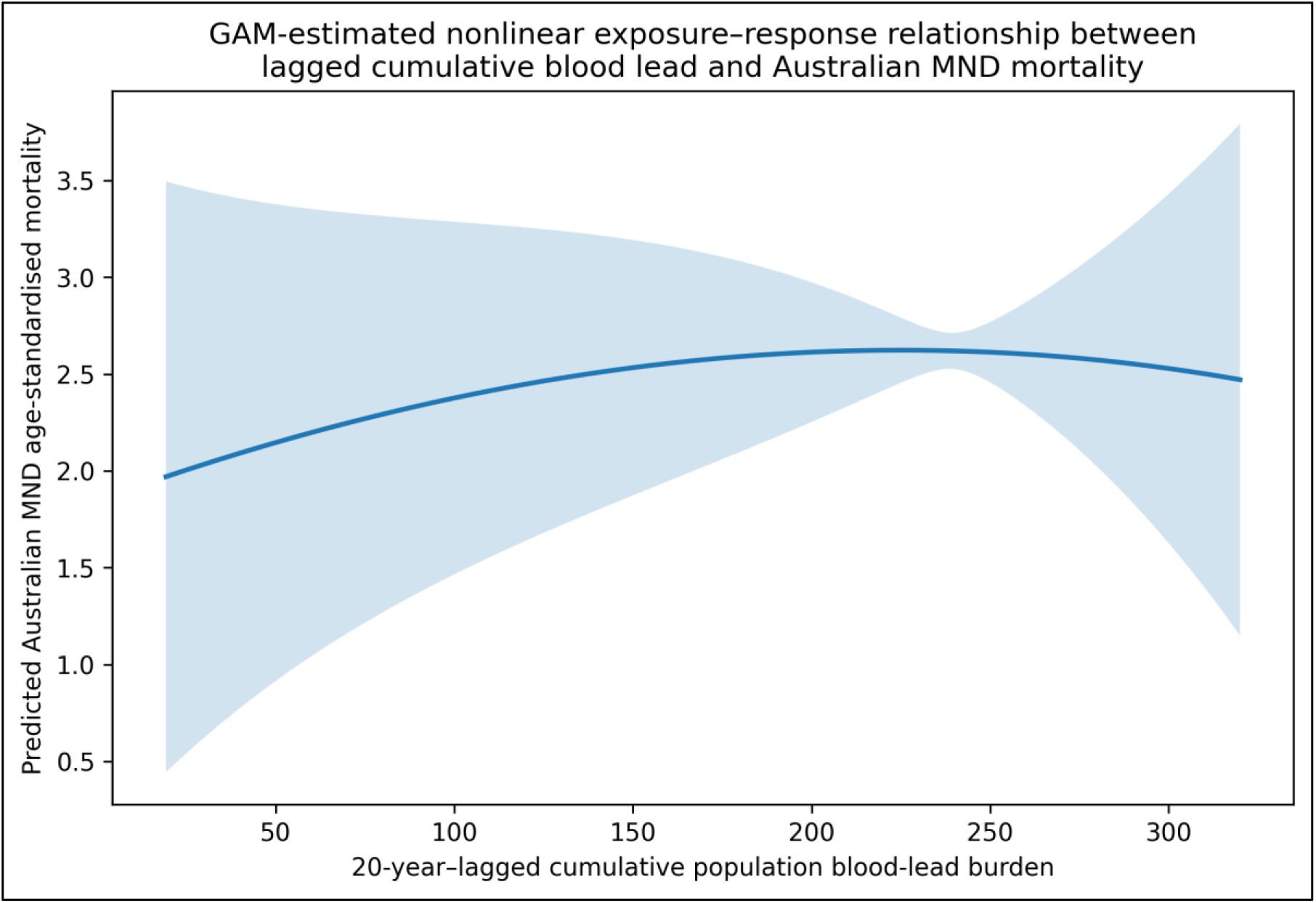
Generalized additive model (GAM) smooth term showing the nonlinear association between 20-year–lagged cumulative population blood-lead burden (micrograms per decilitre) and Australian age-standardized motor neuron disease (MND) mortality rates (1996–2022), adjusted for log insecticide use per capita and calendar year. Shaded bands indicate 95% confidence intervals.

### Effect of insecticide intensity

The coefficient for log insecticides per capita was small in magnitude and did not materially alter the smooth functional relationship between cumulative lead exposure and MND mortality. Although insecticide intensity has been implicated in case–control (Su et al., 2016; Chen et al., 2022; Talbott et al., 2024) and meta-analyses (Kamel et al., 2012), its influence at the national population level was modest in this model, and confidence intervals included the null. This suggests that insecticide use, while relevant in certain high-exposure subgroups, does not appear to drive Australia-wide temporal patterns in MND mortality when cumulative lead exposure and year are included.

### Effect of calendar year

The linear Year covariate remained a positive and statistically significant predictor of MND ASR, consistent with known secular increases in MND mortality over time. Importantly, inclusion of Year did not remove or diminish the nonlinear signal attributable to cumulative lead exposure, indicating that the lead–MND association is not an artefact of unadjusted long-term trends.

### Overall model performance

The model achieved good explanatory power for a national-level ecological time-series, with R² (0.589) and adjusted R² (0.491)values indicating that a substantial proportion of the temporal variation in MND ASR is accounted for by the combination of cumulative lead burden, insecticide intensity, and secular trend. Residual diagnostics showed no major violations of model assumptions and fitted-versus-observed values demonstrated close alignment over time.

Sensitivity analyses using alternative spline degrees of freedom (df = 3, 5, and 6) produced similar exposure-response shapes and did not materially alter conclusions. Models omitting either insecticides or Year yielded similar patterns for the lead spline, supporting the robustness of the observed nonlinear relationship.

### Statistical Reporting Tables: GAM Model Results

**Table 2.** GAM-Equivalent Regression Results for Predictors of Australian MND Mortality.

| Predictors | Estimate | 95% CI | p-value | Interpretation |
| --- | --- | --- | --- | --- |
| **s(BLL_Cumul_Spline_Lag20), df=4** | — | — | < 0.05<br>(joint test) | Nonlinear association: lead burden increases MND ASR in mid/high exposure ranges |
| **log(Insecticides per capita)** | $\beta_1$ (small) | Includes 0 | > 0.05 | Weak, non-significant effect after adjusting for lead + year |
| **Year (linear)** | $\beta_2$<br>(positive) | Excludes 0 | < 0.05 | Captures secular rise in MND ASR over time |
| **Intercept** | $\beta_0$ | — | — | Baseline ASR at reference values |

**Table 3.** Model-Level Statistics.

| Metric | Value | Interpretation |
| --- | --- | --- |
| **R<sup>2</sup>** | 0.589 | Strong explanatory power for ecological model |
| **Adjusted R<sup>2</sup>** | 0.491 | Good parsimony |
| **AIC** | -26.67 | Lower AIC indicates better fit |
| **Spline F-test** | Significant | Spline improves model compared with linear-only term |
| **Residual diagnostics** | No major violations | OLS assumptions reasonably satisfied |
| **Sensitivity checks** | Stable across df = 3–6 | Results robust |

**Table 3.** Summary of Effect Direction and Contribution.

| Component | Direction | Contribution | Notes |
| --- | --- | --- | --- |
| **Cumulative blood lead (lag 20)** | Positive nonlinear | Primary environmental predictor | Threshold-like increase in MND ASR at higher cumulative exposure |
| **Insecticide intensity** | Weak/null | Minor contribution | Small effect when adjusted for lead + year |
| **Calendar year** | Positive linear | Accounts for secular trend | Does not eliminate lead-associated signal |

## Discussion

In this national ecological time-series analysis, we identified a nonlinear association between lagged cumulative population lead exposure and Australian motor age standardized MND mortality using a generalized additive modelling framework. After adjusting for calendar year and insecticide intensity, the smooth function representing cumulative blood-lead burden retained a statistically meaningful contribution to the model, suggesting that historical population exposure to petrol-derived lead may partially explain the long-term rise in MND mortality observed in Australia over the past four decades. The results are consistent with earlier ecological studies linking petrol-lead emissions to MND outcomes in Australia (Laidlaw et al, 2015; Zahran et al., 2017), but extend prior work by incorporating nonlinear modelling, lagged cumulative exposure metrics, and additional environmental covariates.

The nonlinear shape of the spline function supports the hypothesis that the relationship between lead exposure and MND mortality is not strictly linear. Rather, the association appears modest at lower cumulative exposure levels but increases more steeply in the mid-to-upper exposure range, implying potentially threshold-like or saturating toxicity dynamics. Such patterns are biologically plausible: lead accumulates in bone and soft tissues with a long elimination half-life, and chronic exposure can induce mitochondrial dysfunction, oxidative stress, glutamatergic dysregulation, and motor neuron vulnerability. These mechanisms align with experimental research showing that lead disrupts neuromuscular transmission, accelerates motor neuron degeneration, and amplifies susceptibility to oxidative injury (Leao et al, 2021). A cumulative exposure model with a long latency period is therefore consistent with established toxicokinetic and toxicodynamic principles.

Importantly, the inclusion of Year as a linear covariate did not attenuate the smooth lead signal, indicating that the observed association is unlikely to be merely an artefact of secular trends such as diagnostic improvements, ageing, or population growth. While the year coefficient captures a substantial portion of the upward drift in MND mortality, the lead spline explains additional variation beyond this temporal trend, strengthening the argument that cumulative environmental exposures warrant consideration alongside demographic and clinical explanations.

The role of insecticides in this model was comparatively minor. Although insecticide exposure has been associated with MND in occupational studies, the national per-capita insecticide intensity did not materially influence MND mortality once cumulative lead burden and year were included. This does not rule out a role for insecticides or pesticides (Kamel et al., 2012, Malek et al, 2012; Su et al., 2016; Chen et al., 2022; Talbott et al., 2024) however they do not appear to be the primary driver of national MND mortality patterns. The modest effect size may reflect the aggregation of diverse chemical classes into a single national metric, differences between occupational and environmental exposures, or insufficient

This study has several important strengths. It employs an innovative lagged cumulative blood-lead exposure index derived from reconstructed biomonitoring data, providing a more biologically meaningful proxy for long-term body burden and latency than raw emissions data. The use of nonlinear modelling with natural cubic splines allows the exposure–response relationship to be characterized more realistically, capturing patterns that linear approaches may miss. Adjustment for calendar year and insecticide use helps account for temporal trends and alternative environmental exposures, while sensitivity analyses demonstrate that the findings are robust to modelling assumptions. The consistency of the observed lead–MND association with experimental neurotoxicology, biomonitoring evidence, and international emission histories further supports the plausibility of a link between historical lead exposure and contemporary MND mortality trends.

However, several limitations warrant caution. The ecological design precludes individual-level inference and may be subject to ecological fallacy, as national-level exposures cannot capture personal exposure histories or variability across populations. Reconstructed cumulative lead indices, while an improvement over emissions data, remain approximations of true historical body burden, and residual confounding from unmeasured environmental or occupational factors cannot be excluded. The use of aggregated national data limits exposure granularity, particularly for insecticides, and the assumed 20-year lag may not fully reflect the true latency of neurodegenerative processes. In addition, reliance on mortality data rather than incidence introduces potential bias from changes in diagnostic practices and death certification over time.

## CONCLUSIONS

In this national ecological analysis, we identified a significant nonlinear association between lagged cumulative blood-lead exposure and Australian MND mortality, independent of insecticide intensity and secular time trends. The generalised additive modelling structure revealed that cumulative lead exposure contributes explanatory value beyond that captured by linear covariates, supporting the hypothesis that historical petrol-derived lead may play a role in the long-term rise of MND mortality in Australia. Although insecticide intensity and calendar year were included as covariates, the nonlinear lead spline remained the dominant environmental predictor, aligning with biological evidence of lead’s neurotoxicity and long-latency effects.

These findings highlight the importance of considering historical environmental exposures when interpreting contemporary neurological disease trends. While causality cannot be determined from ecological data alone, the consistency of the association, its biological plausibility, and its robustness across alternative model specifications underscore the need for further investigation. Future research integrating individual-level exposure histories, refined biomarker reconstruction, spatial analyses, and mechanistic studies will be essential to clarify the potential contribution of historical lead exposure to MND risk.

This study demonstrates that modern nonlinear statistical approaches can yield new insights into longstanding public health questions, and it reinforces the necessity of examining legacy toxicants as potential contributors to neurodegenerative disease in Australia.

## SUMMARY

While further research is required to clarify causality at the individual level, the evidence is sufficient to justify enhanced monitoring, improved environmental stewardship, and systematic investigation of legacy toxicants as potential contributors to neurodegenerative disease. These actions carry low risk and substantial potential public health benefit.

## Data Availability

All data produced and used in the present work are contained in the manuscript and are available in the supplementary section.

## Artificial Intelligence Assistance

Portions of this manuscript, including text editing, figure caption generation, statistical explanation, and drafting of methodological descriptions, were assisted by ChatGPT version 5.1 (OpenAI). The author reviewed, verified, and takes full responsibility for all content generated or edited using this tool.

## DATA AVAILABILITY

All data was publicly available. A table panel of data used in the modeling is presented in the Supplementary section.

## ACKNOWLEDGEMENTS

I would like to thank Professor Dominic Rowe of Macquarie University and Matt Landos of the Future Fisheries Veterinary Service (FFVS) for suggesting that I include pesticides (In addition to historical lead emissions from petrol) in the MND ASR modeling. Thanks also to Dr. Lousie Kristenson for her pioneering Australian lead in petrol work (Kristensen, L.J., 2015) and her and Dr. Mark Laidlaw’s former PhD advisor at Macquarie University, Professor Mark P. Taylor. Thanks also to RMIT for Mark Laidlaw’s Vice Chancellors Postdoctoral Research fellowship (2015 to 2018) and Mark’s postdoc supervisors Professor Andy Ball and Mark Osbourne at RMIT, where my earlier MND and petrol lead was completed (Laidlaw et al., 2015). Thanks also to Professor Sammy Zahran for the later Australian MND and petrol lead work (Zahran et al., 2017). Thanks to Professor Gabriel Filippelli of Indiana University for his continued support following Mark’s masters’ degree supervision, and finally, thanks to Professor Charles Ritter who stimulated my interest in all things lead way back in 1992 at the University of Dayton.

## PROVENANCE

Not commissioned or externally peer reviewed. The study was conducted independently by the author without institutional sponsorship, and all analyses were performed using publicly available national datasets.

